# Human in vivo immunology of tuberculosis is not affected by sex dimorphism

**DOI:** 10.64898/2026.07.20.26358462

**Authors:** Jana Jiang, James Greenan-Barrett, Rishi K Gupta, Mahdad Noursadeghi, Carolin T Turner

## Abstract

Males incur greater risk of tuberculosis (TB) than females, but the contribution of sex-associated immune differences remains unclear. We addressed this using sex-stratified transcriptomic analyses across four independent studies spanning active pulmonary TB, subclinical TB and latent infection, in peripheral blood, bronchoalveolar lavage (BAL) and by using the tuberculin skin test (TST) as a standardised in vivo antigenic challenge. In blood of active TB patients, expression of TNF- and type I interferon-regulated signatures, genome-wide gene expression, and performance of leading host-response biomarkers of TB were comparable between sexes. Similarly, blood transcriptomic biomarkers showed no meaningful sex-related differences for predicting asymptomatic or incident TB. In the TST of people with latent infection, bulk and single-cell RNA sequencing identified only limited differences, largely restricted to sex chromosome-linked transcripts, with no consistent evidence of dimorphism in immune-regulated pathways. Single-cell RNA sequencing of BAL samples identified reduced abundance of B cells in male TB patients, with gene expression differences again largely restricted to sex chromosome-linked transcripts. These findings suggest that canonical immune responses associated with TB are broadly similar between the sexes, and that increased TB risk among males more likely reflects differential exposure rather than intrinsic immunological susceptibility.

## Introduction

Tuberculosis (TB) is a destructive chronic inflammatory disease caused by infection with *Mycobacterium tuberculosis* (*Mtb*). Males are at greater risk of TB disease than females, with a male-to-female incidence ratio of approximately 1.6.^1^ Prevalence surveys suggest that this gap is not explained by differences in care-seeking or reporting alone.^2,3^ Males with TB have a higher bacillary burden and experience worse clinical outcomes including higher mortality, suggesting some degree of differential biological susceptibility to disease.^4,5^ However, population-based *Mtb* immunoreactivity surveys show males experience greater incident infections after puberty,^6^ but similar rates of disease progression to females,^7^ indicating that sex-associated differences in exposure^8,9^ are more important determinants of the male bias in TB than sex-associated differences in immune responses. The extent to which differential disease-risk is determined by sexual dimorphism in immune responses or by social and behavioural determinants remains unclear.

The genetic basis of sex-biased immunity arises partly from the asymmetry of the X and Y chromosomes. The X chromosome contains approximately 800 protein-coding genes, whereas the Y chromosome contains fewer than 80, leading to potential differential gene-dosage effects.^9,10^ Although dosage compensation is usually achieved through X-chromosome inactivation (XCI),^11,12^ approximately 15-25% of X-linked genes escape XCI.^13,14^ A key example is TLR7, which exhibits biallelic expression in a proportion of female immune cells with potential to increase innate immune sensing upstream of type 1 IFN responses.^15^ In addition, gonadal sex hormones are reported to have wide-ranging immunomodulatory effects.^16^ In particular, oestradiols increase IFNα production by plasmacytoid dendritic cells (pDCs),^17–19^ whereas testosterones suppress type I IFN signalling in both pDCs and monocytes, and enhance pro-inflammatory TNF responses.^20^ Type I IFN and TNF signalling are evident in the immunology of TB and can contribute to both host-protection and disease pathogenesis. Indeed, a recent mouse study that uncoupled gonadal from chromosomal sex by translocating the *Sry* (sex-determining region Y) locus from the Y chromosome to an autosome found gonadal sex to be the primary driver of male susceptibility and female resistance to TB.^21^ This was accompanied by increased myeloid responses and TNF levels in male lungs, and increased adaptive responses, including elevated CD4 Th1 cells and IFNg producing B cells in female lungs.^21^ However, direct evidence for sex-associated differential activity of these immunological pathways in humans is lacking.

In the present study, we sought to address this knowledge gap by sex stratified analysis of transcriptional perturbation in TB blood and bronchoalveolar lavage (BAL) samples, and molecular profiling of the tuberculin skin test (TST) as a standardised human experimental challenge to model in vivo immunity in TB. We tested the hypothesis that TNF responses are increased and type 1 IFN responses attenuated in males compared to females and explored the possibility of sexual dimorphism in IFNg/Th1 signalling that is also known to be a determinant of outcomes in *Mtb* infection.

## Materials and Methods

### Study approvals

All datasets analysed in this study were obtained from previously published studies, each of which had received ethical approval as described in the original publications.^22–26^ No additional ethical approval was required for this secondary analysis.

### Study populations

Data from four published studies were analysed: First, the active TB blood cohort comprised bulk RNA sequencing data from a prospective diagnostic accuracy study of 181 consecutive adults presenting for investigation of pulmonary TB in Cape Town, South Africa, of whom 53 had microbiologically confirmed disease (27 men, 26 women).^22^ Second, the subclinical TB cohort included individual participant data from a recent meta-analysis of blood RNA signatures for subclinical TB.^23^ Of the seven included datasets, six had sex data available: four RNA sequencing datasets (the Adolescent Cohort Study (ACS) of IGRA- or TST-positive individuals from South Africa^27^, the Grand Challenges 6-74 (GC6-74) study of household contacts from South Africa, The Gambia and Ethiopia^28^, and two close contact studies from London^29^ and Leicester^30^ and two qPCR datasets of individuals from TB-endemic communities in South Africa (CORTIS-01^31^ and CORTIS-HR^32^). Together these comprised 3,817 participants (2,137 women, 1,675 men, 5 unknown sex), of whom 154 (83 women, 71 men, none of unknown sex) progressed to microbiologically confirmed TB disease over a 12-month interval.^23^ Third, the latent TB cohort included RNA sequencing data from tuberculin skin test (TST) samples.^24,25^ This cohort comprised healthy adults aged 18-60 with evidence of *Mtb*-specific immune memory by IFNg release assay but no clinical or radiological features of active disease.^24,25^ Participants received intradermal injections of tuberculin or saline, with punch biopsies taken at day 2 and day 7 post-injection, reflecting maximum clinical inflammation and maximum T cell infiltration, respectively.^33^ TST biopsies were available from 216 participants at day 2 (115 women, 101 men) and 158 at day 7 (85 women, 73 men). Saline control biopsies were obtained from a separate group of 33 volunteers (19 women, 14 men). Lastly, negative pressure was applied to the skin overlying TST injection sites to induce suction blisters, and the resulting blister fluid was harvested for single-cell RNA sequencing. Single cell data from day 2 samples were available from 31 participants (22 women, 9 men). Fourth, the active TB BAL cohort comprised single cell RNA sequencing data of 21 adults (11 women, 10 men) with pulmonary TB in Leicester, UK, diagnosed based on clinical signs and symptoms and imaging, and supported by either microbiological confirmation or treatment response.^26^

### Data processing

Gene expression count matrices and processed data objects were obtained directly from the original studies and used without reprocessing.^22–26^

### Transcriptional modules

Transcriptional modules represent cytokine-inducible gene expression in a specific cell type or tissue context. Blood, keratinocyte (KC) and monocyte-derived macrophage (MDM) modules have previously been derived and validated. Blood modules include the STAT1-regulated module, representing type 1 IFN responses,^34^and the TNF-inducible module, representing TNF responses.^35^ KC modules include IFNA-, IFNG- and TNF-inducible genes that were specifically upregulated in KC cultures in response to cognate cytokine.^36^ MDM modules include TNF-inducible genes that were specifically upregulated in MDM cultures after 24 hr stimulation with cognate cytokine compared to other cytokines, as well as IFN1- and IFN2-inducible genes that were specifically upregulated after 4 hr stimulation in response to one of the interferon stimuli but not the other.^37^ TST modules were identified previously through upstream regulator analysis of the TST response transcriptome.^24^ The gene lists that make up each module are listed in Supplementary Table 1.

Module scores were calculated as the arithmetic mean log_2_ expression of constituent genes per sample (for bulk RNAseq data) or per cell (for single-cell RNAseq data). Statistical differences in module scores between men and women were assessed with unpaired, two-sided Wilcoxon tests and corrected for multiple testing using False Discovery Rate (FDR). FDR-adjusted p values <0.05 were considered significant.

### Diagnostic signature evaluation

In the active TB blood cohort, the four blood transcriptional diagnostic signatures with the highest diagnostic accuracy in a prospective head-to-head comparison of 27 candidate signatures^22^ were compared between the sexes: BATF2,^38^ Kaforou25,^39^ Roe3^29^ and Sweeney3.^40^ These signatures met or approximated the minimum WHO target product profile for a TB triage test independent of age, sex, HIV status, previous TB or sputum smear result.^22^ In the subclinical TB cohort, discriminative performance was assessed using area under the receiver operating characteristic curves (AUROCs) with 95% confidence intervals, estimated using DeLong’s method for the five best performing single-gene transcripts (ANKRD22, BATF2, FCGR1A/B, GBP2 and SERPING1) and the three best performing multi-gene signatures (Darboe11,^41^ PennNicholson6^42^ and Roe3^29^). Analyses were stratified by sex and restricted to participants with available sex information. Differences in AUROCs between males and females were tested using unpaired DeLong’s test, with p-values adjusted for multiple testing using the Benjamini-Hochberg method.

### Bulk RNAseq differential gene expression analysis

Genome-wide differential expression analysis was performed using DESeq2 (v1.44.0) via the SARTools pipeline (v1.8.2) in R (4.4.0). In the active TB blood cohort, differential expression between the sexes was assessed using thresholds of FDR < 0.05 and absolute log_2_ fold difference ≥ 1 (fold change ≥ 2). In the TST cohort, differential expression was performed within the TST-response transcriptome, defined as upregulated differential gene expression relative to saline controls,^25^ using the same thresholds.

### Single-cell RNAseq analysis of TST blisters and BAL samples

Single-cell sequencing data from TST blisters were processed and annotated as described.^25^ The dataset comprised 63881 cells from n=31 Day 2 TST blisters (n=22 female, n=9 male) and was clustered into 101 discrete clusters, which were collapsed into 14 distinct cell types with one cluster remaining undefined. The undefined and erythrocyte clusters were excluded from sex-stratified analysis.

A processed and annotated single-cell sequencing dataset of BAL samples was downloaded from Gene Expression Omnibus, accession number GSE326212. Only samples from active TB patients were retained, resulting in 241,899 cells from n=21 patients (n=11 female, n=10 male), distinguished into 27 distinct cell types as described in the original publication.^26^

#### Differential abundance analysis

For comparison of cell type or cluster abundance between samples from men and women, the number of cells belonging to each cell type or cluster were summed for each sample. The R package edgeR was used for differential abundance analysis, employing first the filterByExpr function with default settings to remove low abundance cell types/clusters, then the estimateDisp function (with trend=”none”) and glmQLFit function (with robust=TRUE and abundance.trend=FALSE) to implement a negative binomial generalised linear model with quasi-likelihood F (GLM-QLF) test. Cell types or clusters with FDR <0.05 were considered differentially abundant.

#### Calculation of module scores

The module score for each cell was calculated as the arithmetic mean expression of the constituent module genes, using log_2_-normalised gene counts, including signature genes with zero expression. Module scores for each cell were Z-score scaled compared to all cells in the dataset and averaged for each participant and cell type.

#### Single-cell RNAseq differential gene expression analysis

For comparison of gene expression between men and women at cell type level, the aggregateAcrossCells function from R’s scuttle package was used to create pseudobulk profiles by summing raw counts for each gene across cells belonging to the same cell type and sample. Pseudobulk profiles from less than 20 cells were filtered out before statistical analysis was performed. The number of sex-stratified pseudobulk datasets per cell type are listed in Supplementary Table 2). Differentially expressed genes were identified using the pseudoBulkDGE function from R’s scran package, with method = “edgeR”. Genes with FDR <0.05 were considered significant.

### Data and code availability

All data analysed during this study are available from public repositories as listed in Supplementary Table 5. The processed data objects used for sex-stratified analyses are available from UCL’s Research Data Repository at https://doi.org/10.5522/04/33026150 upon peer-reviewed publication of this manuscript. Analysis code is available at https://github.com/carolinturner/sexual_dimorphism_tb. A source data file is provided for all figures.

## Results

### Sex-stratified blood transcriptional perturbation in TB

Blood transcriptional perturbation in TB is widely reported. We have previously identified blood transcriptional signatures attributable to type 1 IFN (via STAT1 signalling)^34^ and TNF signalling.^35^ We compared the expression of these signatures in bulk RNA sequencing data from men (n = 27) and women (n= 26) with microbiologically confirmed pulmonary TB in Cape Town, South Africa.^22^ Neither the STAT1 nor the TNF signature expression differed between male and female TB patients (Fig. 1a). Likewise, alternative stimulus-specific modules of type 1 IFN, type 2 IFN and TNF-inducible gene expression, derived from monocyte-derived macrophages,^37^ were comparable between sexes in these data (Supplementary Fig. 1).

**Figure 1.**
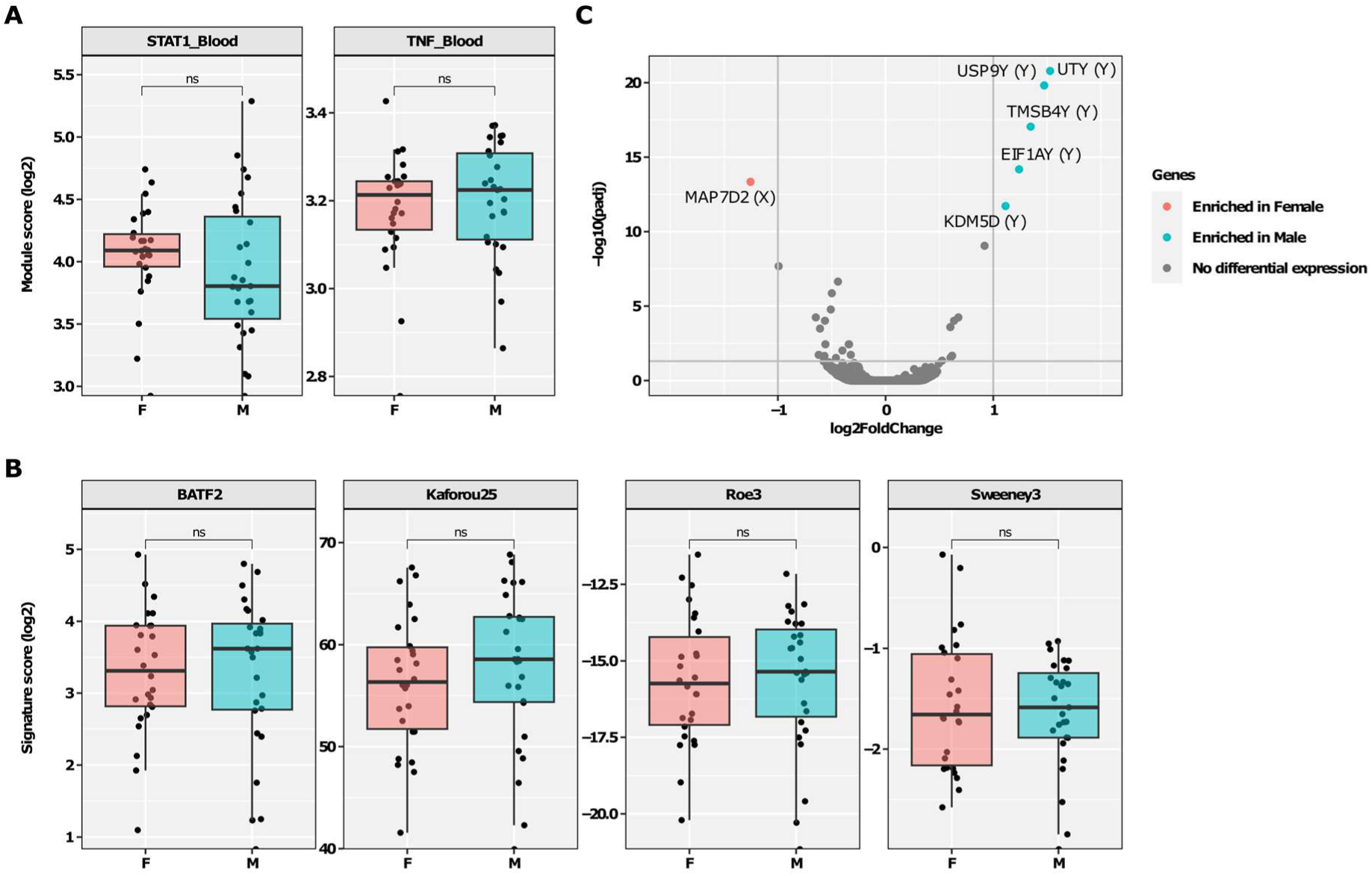
No sex differences in blood immune perturbations during active tuberculosis (TB). Bulk RNA sequencing data from whole blood samples of patients with active TB disease were compared between male (M, n=27) and female (F, n=26) participants. **A.** Expression of STAT1-regulated and TNF-inducible transcriptional modules as proxies for type I interferon and TNF cytokine activity in blood, respectively. **B.** Expression of four transcriptional gene signatures for the diagnosis of active TB. Boxplots in **A-B** depict median and inter-quartile range and include individual data points. Statistical significance was assessed with unpaired, two-sided Wilcoxon tests (ns p>0.05). **C.** Volcano plot showing statistical significance against quantitative gene expression differences between male and female blood. Genes highlighted in red and turquoise are upregulated (FDR<0.05 and fold change ≥2) in female and male, respectively, and are labelled with gene name. Chromosome origin is indicated in brackets.

Blood transcriptional perturbation in TB has been widely used to identify a range of TB host response biomarkers downstream of TNF and IFN signalling. We had previously reported equivalent diagnostic accuracy for these biomarkers between male and female sex.^22^ In the present analysis we also found that none of the four best performing biomarkers (BATF2,^38^ Kaforou25,^39^ Roe3^29^ and Sweeney3^40^) showed sex-stratified differences in expression within this clinical cohort (Fig. 1b). TB-associated blood transcriptional biomarkers have also been shown to be enriched prior to the diagnosis of TB, likely reflecting host responses in subclinical TB. In data from a previously published individual participant level meta-analysis including 3,817 people,^23^ we found comparable diagnostic accuracy between male and female sex for the best performing biomarkers for incident TB (Table 1). AUROCs ranged from 0.71 (95% CI 0.64-0.79) to 0.78 (95% CI 0.72-0.84) in women and from 0.76 (95% CI 0.7-0.82) to 0.81 (95% CI 0.75-0.86) in men.

**Table 1.** Discriminative performance of blood transcriptional signatures for subclinical TB, stratified by sex. Area under the receiver operating characteristic curve (AUROC) with 95% confidence intervals for the five best-performing single-gene transcripts (ANKRD22, BATF2, FCGR1A/B, GBP2 and SEPING1) and three multi-gene signatures (Darboe11, PennNicholson6 and Roe3), estimated using individual participant data from four cohorts (n=3,817). P-values for sex differences were calculated using unpaired DeLong’s test and adjusted for multiple testing using the Benjamini-Hochberg method.

| Gene Signature | AUC Female (95% CI) | AUC Male (95% CI) | Adjusted p-values |
| --- | --- | --- | --- |
| ANKRD22 | 0.78 (0.72 - 0.84) | 0.79 (0.71 - 0.87) | 0.812 |
| BATF2 | 0.76 (0.7 - 0.81) | 0.78 (0.72 - 0.84) | 0.810 |
| Darboe11 | 0.71 (0.64 - 0.79) | 0.76 (0.7 - 0.82) | 0.712 |
| FCGR1A/B | 0.73 (0.68 - 0.79) | 0.81 (0.75 - 0.86) | 0.586 |
| GBP2 | 0.74 (0.68 - 0.79) | 0.78 (0.72 - 0.84) | 0.712 |
| PennNicholson6 | 0.73 (0.65 - 0.81) | 0.78 (0.72 - 0.84) | 0.712 |
| Roe3 | 0.75 (0.7 - 0.81) | 0.78 (0.73 - 0.84) | 0.749 |
| SERPING1 | 0.75 (0.69 - 0.8) | 0.76 (0.7 - 0.82) | 0.812 |

Finally, we explored genome-wide differential gene expression analysis between male and female patients in the active TB disease blood cohort. We identified only 5 transcripts that were enriched in males (KDM5D, USP9Y, TMSB4Y, UTY and EIF1AY), all encoded on the Y chromosome, and 1 gene that was enriched in females (MAP7D2), encoded on the X chromosome (Fig. 1c). No immune responses genes were differentially expressed. Taken together, we found no sex-associated differences in the immunology of TB reflected in blood transcriptional perturbation.

### Sex-stratified immune responses at the site of the TST

Next, we sought to evaluate sex differences in human in vivo immune responses at two time points (day 2 and day 7) after a standardised antigenic challenge by bulk RNA sequencing of TST tissue biopsies from previously immune sensitised people without evidence of TB disease.^24^ The day 2 time point in the TST reflects peak inflammatory responses, whereas the day 7 time point reflects local evolution of the *Mtb*-reactive T cell repertoire.^24^ Within these reactions, we first focussed on selected cytokine-inducible activity by comparison of cytokine-specific gene expression signatures derived independently from keratinocytes (KC) stimulated with each of IFNα, IFNg and TNF.^36^ The expression of KC-derived modules for each of these cytokines showed no significant differences between male and female sex at either day 2 or day 7 (Fig. 2a). Likewise, we found no sex-associated differences in IFN- or TNF-inducible gene expression signatures derived from the TST transcriptomic data itself (Fig. 2b),^24^ or in previously published cytokine-inducible transcriptional signatures derived from MDM ± selected cytokine stimuli^37^ (Supplementary Fig. 2). Genome-wide differential gene expression between male and female sex revealed only one transcript, for the chemokine CXCL8, to be statistically significantly enriched in males in the day 2 TST and no statistically significant differences in the day 7 TST (Fig. 2c-d).

**Figure 2.**
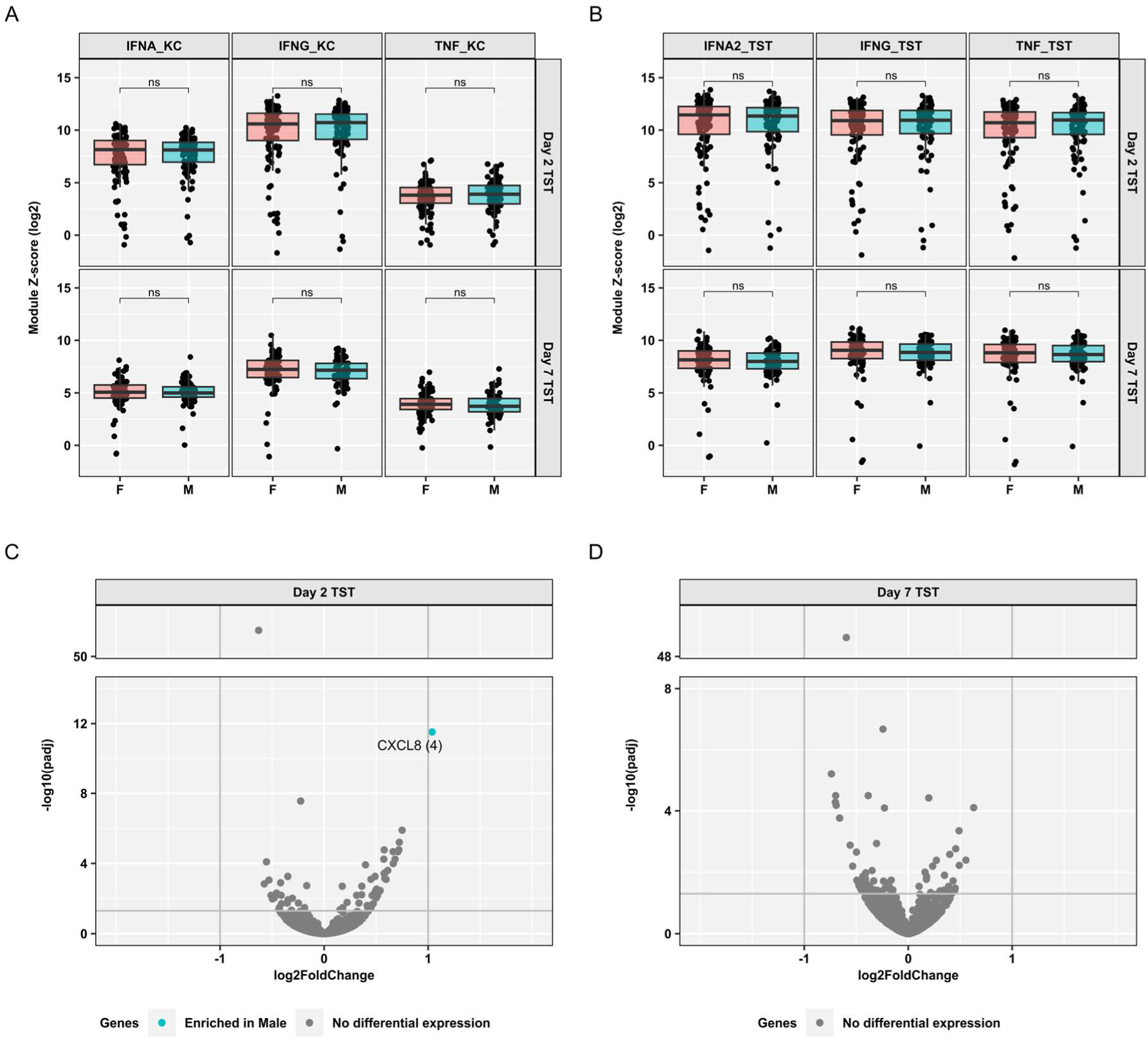
No sex differences in tuberculin skin test (TST) bulk immune responses during latent TB. Bulk RNA sequencing data from TST samples of people with latent TB were compared between male (M) and female (F) participants on day 2 (n=101 male, n=115 female) and day 7 (n=73 male, n=85 female) following TST challenge. **A-B.** Expression of interferon- and TNF-inducible transcriptional modules in Day 2 and Day 7 TSTs, shown as Z-score scaled TPM expression using saline-challenged skin biopsies from a separate cohort of participants as control group. Modules reflect cytokine activity in keratinocytes (KC modules; panel **A**) or co-correlated gene expression changes in the TST that are bioinformatically predicted to be regulated by an upstream cytokine (TST modules; panel **B.**). Boxplots depict median and inter-quartile range and include individual data points. Statistical significance was assessed with unpaired, two-sided Wilcoxon tests (ns p>0.05). **C-D.** Volcano plots showing statistical significance against quantitative gene expression differences between male and female TST samples, taken on Day 2 (**C.**) or on Day 7 (**D.**). The gene highlighted in turquoise in (**C.**) is upregulated (FDR<0.05 and fold change ≥2) in male participants and labelled with gene name. Chromosome origin is indicated in brackets.

Finally, we investigated the possibility that sex-associated differences in TST immune responses may be masked in bulk transcriptional data due to cell-type specific effects. We addressed this by analysis of single-cell RNA sequencing data from TST skin blisters in 9 males and 22 females. The integrated dataset contained 63881 cells and was clustered into 101 discrete clusters which were collapsed into 13 distinct cell types.^25^ Differential abundance analysis identified basal keratinocytes as more abundant in females (FDR = 0.01), with no other cell types reaching significance (Supplementary Table 3). At the cluster level, significant differences were similarly limited to basal keratinocyte clusters being more abundant in females (FDR = 0.04). Next, we compared cytokine-regulated module scores for IFNα, IFNg and TNF across cell types. TST-derived module scores were comparable between men and women (Fig. 3a). Equally, keratinocyte-derived and MDM stimulus-specific modules for IFNα, IFNg and TNF were equivalent between the sexes across all cell types (Supplementary Fig. 3a-b). Pseudobulk differential gene expression analysis revealed 31 differentially expressed genes between male and female sex across discrete cell subsets (FDR < 0.05), of which 14 were enriched in males (10 Y chromosome-linked, 4 autosomal) and 17 in females (all X chromosome-linked) (Fig. 3b). The enrichment of four autosomal genes in men was limited to single cell types and included HBB (hemoglobin subunit B) in myeloid cells, FES (FES Proto-Oncogene, Tyrosine Kinase) in NK cells, KIR3DL2 (an inhibitory killer cell immunoglobulin like receptor) in γδ V1 T cells and the pseudogene SLC35E2A in CD8 T cells. None of these genes is commonly known for increased expression in men. Of the 17 transcripts enriched in women, two are non-coding RNAs that regulate X-chromosome inactivation (XIST and JPX)^43^ and 14 are protein-coding X-linked genes known to escape XCI (EIF1AX, ZRSR2, ZFX, EIF2S3, SMC1A, KDM6A, KDM5C, CXorf38, TXLNG, PRKX, ARSD, PUDP, RPS4X and SEPTIN6).^14,44–47^ Similarly, PPP1R2C has been reported to be down-expressed in Turner syndrome patients who lack a second X chromosome compared to healthy females.^48^ This pattern of predominantly sex-chromosome-linked differential expression was evident across individual cell types (Fig. 3b).

**Figure 3.**
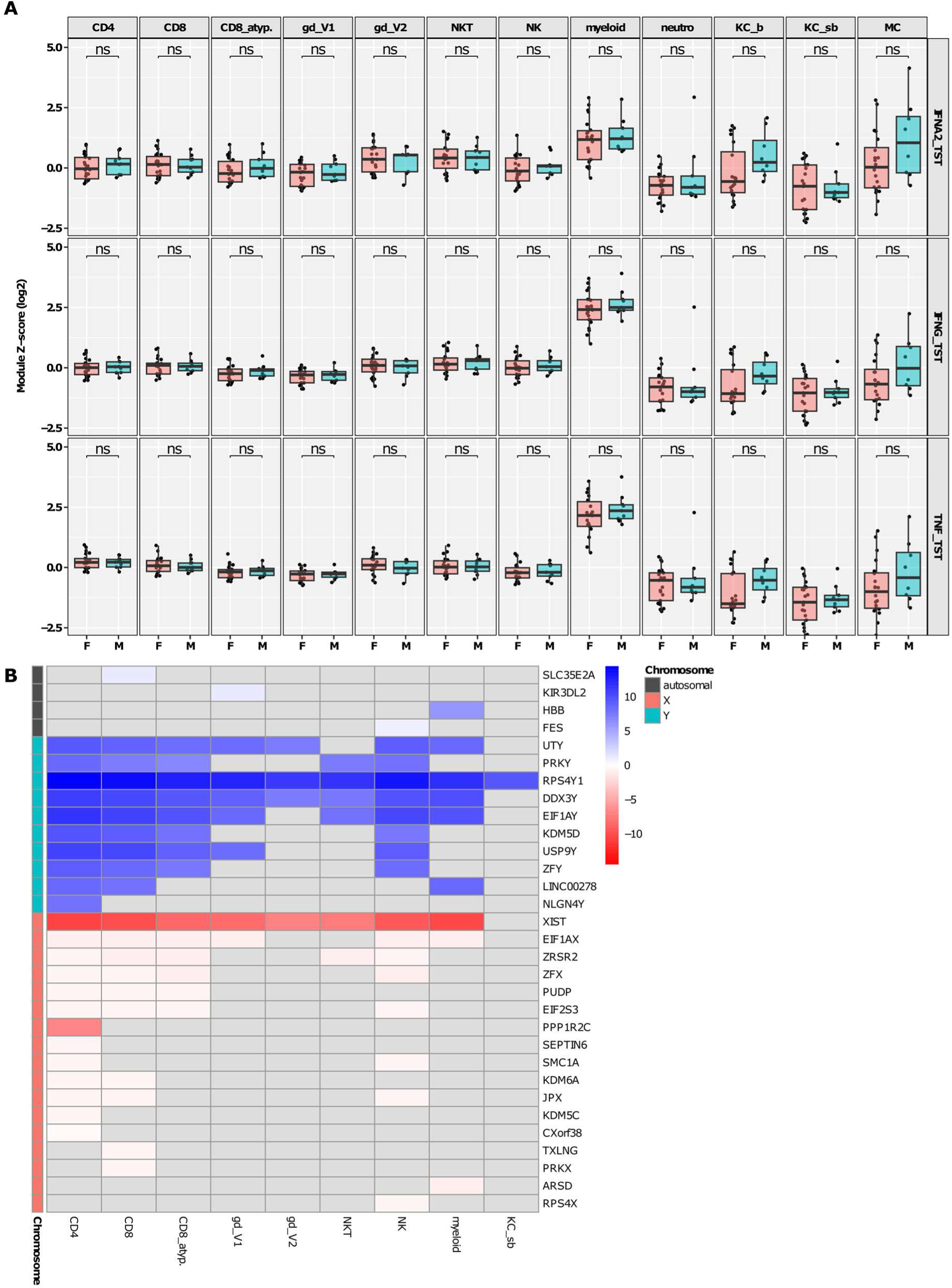
No sex differences in tuberculin skin test (TST) single-cell immune responses during latent TB. Single-cell RNA sequencing data from Day 2 TST blisters were compared between male (M, n=9) and female (F, n=22) participants. Cell type annotations include CD4 T cells (CD4), conventional CD8 T cells (CD8), atypical CD8 T cells (CD8_atyp., characterised by reduced expression of CD8B and alpha/beta TCRs compared to conventional CD8s), Vδ1 (gd_V1) and Vδ2 (gd_V2) gamma-delta T cells, natural killer T cells (NKT), natural killer cells (NK), myeloid cells (myeloid), neutrophils (neutro), basal (KC_b) and suprabasal (KC_sb) keratinocytes and melanocytes (MC). **A.** Expression of interferon- and TNF-inducible transcriptional modules in Day 2 TST suction blister cells. Modules reflect co-correlated gene expression changes in the TST that are bioinformatically predicted to be induced by an upstream cytokine. Boxplots depict median and inter-quartile range and include individual data points. Statistical significance was assessed with unpaired, two-sided Wilcoxon tests, adjusted for multiple testing (ns FDR>0.05). **B.** Heatmap of differentially expressed genes (rows; FDR<0.05) between male and female TST blister samples, stratified by cell type (columns) and chromosome origin (annotation colour). Cell types without any differentially expressed genes between sexes are not included. Heatmap colour range indicates the log2 fold change between men and women, showing genes that are enriched in men in blue, genes that are enriched in women in red, and genes with FDR>0.05 in grey.

### Sex-stratified immune responses in BAL samples in TB

A limitation of the Day 2 TST blister model is the absence of B cells and low abundance of neutrophils (<1% of all blister cells). Therefore, we also evaluated sex differences in BAL samples from active TB patients,^26^ which contain a broad range of lung immune cell populations relevant to TB pathogenesis, including alveolar macrophages, T cells, B cells and neutrophils. Differential cell type abundance analysis of a dataset of 241,899 BAL cells from 11 females and 10 males identified B cells as less abundant in males (Fig. 4a; logFC = -2.09, FDR 0.015), with none of the other 26 cell types reaching significance (Supplementary Table 4). Cytokine-regulated MDM transcriptional modules were comparable between men and women across all cell types, with no significant differences after multiple testing correction (Fig. 4b). Pseudobulk differential gene expression analysis found 9 genes enriched in males (Fig. 4c), of which 7 were Y chromosome-linked whilst the 2 autosomal genes (DDX43 and COL15A1) have previously been described to show male-biased expression.^49,50^ Consistent with our findings in the TST model, this tendency for sex-chromosome–linked differential expression was observed across individual cell types (Fig. 4c).

**Figure 4.**
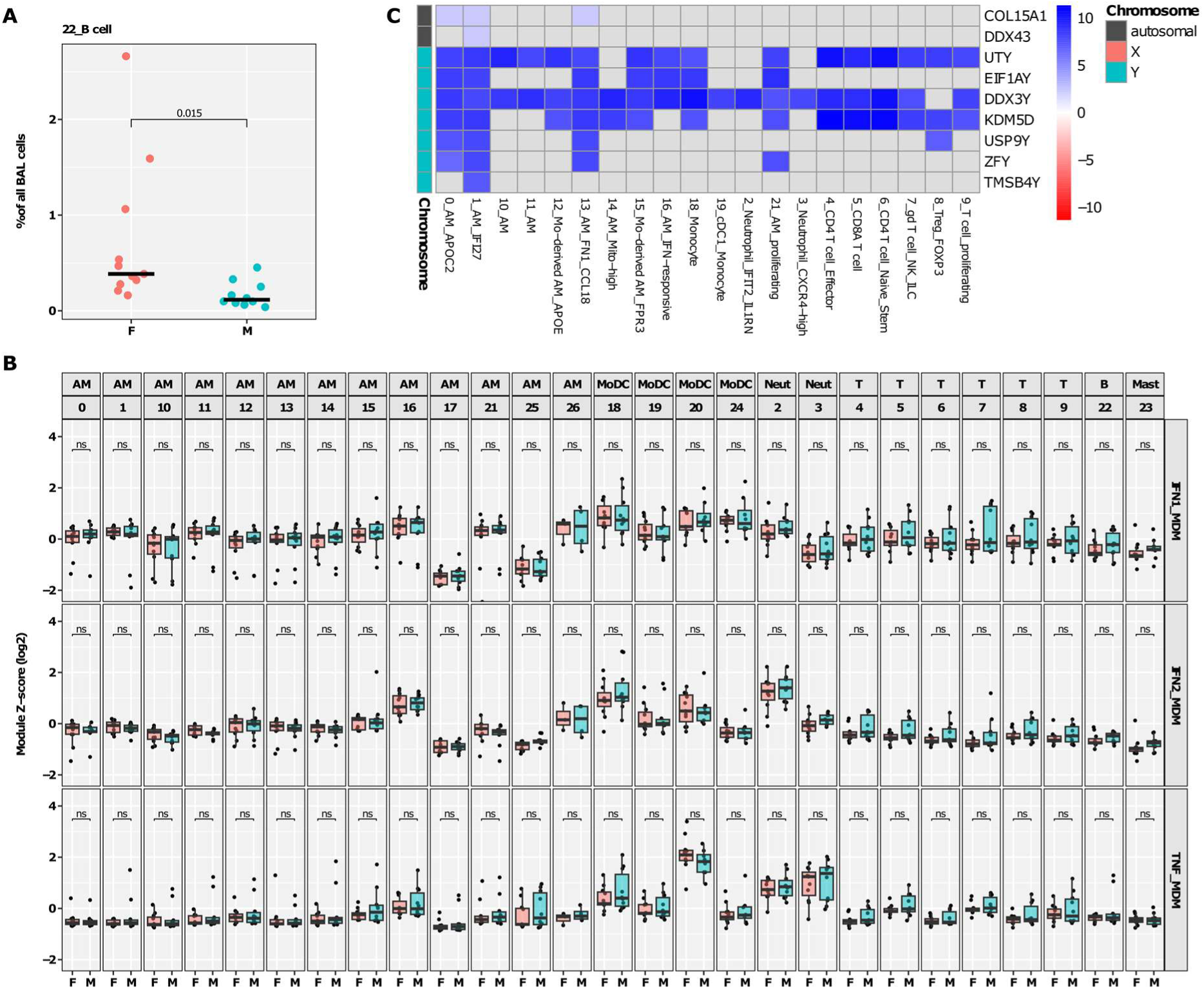
Fewer B cells in bronchoalveoloar lavage (BAL) from male TB patients. Single-cell RNA sequencing data from BAL samples were compared between male (M, n=10) and female (F, n=11) TB patients. Clustering annotations included 27 cell types, encompassing subsets of alveolar macrophages (AM), monocytes (Mo), dendritic cells (DC), neutrophils (Neut), mast cells (Mast), B and T cells. **A.** B cell abundance by sex, shown as percentage of BAL cells, with FDR as determined by differential abundance analysis with edgeR (Supplementary Table 4). **B.** Expression of interferon- and TNF-inducible transcriptional modules in BAL cells, stratified by broad cell type category and cluster number. Module scores were first calculated for each cell, then Z-score scaled compared to all cells in the dataset and finally averaged for each participant and cell type. Modules reflect cytokine activity in monocyte-derived macrophages (MDM), derived from stimulant-specific gene expression changes after in vitro MDM stimulation. Boxplots depict median and inter-quartile range and include individual data points. Statistical significance was assessed with unpaired, two-sided Wilcoxon tests, adjusted for multiple testing (ns FDR>0.05). **C.** Heatmap of differentially expressed genes (rows; FDR<0.05) between male and female BAL samples, stratified by cell type (columns) and chromosome origin (annotation colour). Cell types without any differentially expressed genes between sexes are not included. Heatmap colour range indicates the log2 fold change between men and women, showing genes that are enriched in men in blue, and genes with FDR>0.05 in grey.

## Discussion

In this study we tested the hypothesis that male and female sex exhibit differences in TNF and IFN mediated immune responses to tuberculosis (TB) across four independent studies spanning the spectrum of TB disease states and immunological compartments. We found no evidence of sexual dimorphism in the in vivo human cell-mediated immunology of TB.

A key strength of the study is the application of independently derived gene expression signatures that provide a measure of the functional activity of selected immune pathways of interest. We evaluated these pathways both in steady state peripheral blood transcriptomic perturbation associated with both subclinical and symptomatic TB disease, and in a dynamic response to a standardised time-indexed immunogenic stimulus using the TST. Bulk transcriptomic analysis in blood and TST is particularly valuable to quantify multicellular immunological signalling at systems level, but cannot assess differential immune responses associated with sex at the level of individual cell types. We addressed this limitation by our additional analysis of single cell transcriptomic data from TST blisters. Of note, TST single cell data did not include neutrophils or B cells, which therefore represent specific immunological components we were unable to evaluate in this context. However, TST blister samples did include monocyte derived and T cell populations which are generally thought to be most important in TB.^24,25^ Finally, we leveraged recently published single cell data from bronchoalveolar lavage samples in active TB, which contain neutrophils and B cells. In this analysis, we found reduced B cell numbers in male patients, consistent with previously reported smaller B cell follicles in *Mtb*-infected mice.^21,51^

In addition to the targeted evaluation of TNF and IFN-regulated immune response pathways, we broadened our analysis of sex-associated differences in immune responses using genome-wide differential gene expression analysis. This too, failed to identify any robust differences in the activity of immunological pathways. Importantly, this analysis did reveal examples of differential gene expression between sexes encoded by sex chromosomes confirming that we had adequate statistical power to identify statistically significant differences between male and female sex.

Taken together, we conclude that the canonical human cell-mediated immunology associated with TB in vivo is not affected by sex, and that increased epidemiological risk of *Mtb* infection and incident TB disease in post pubertal males most likely arises from increased behavioural exposure risk.

## Supplementary Figures

**Supplementary Figure 1.**
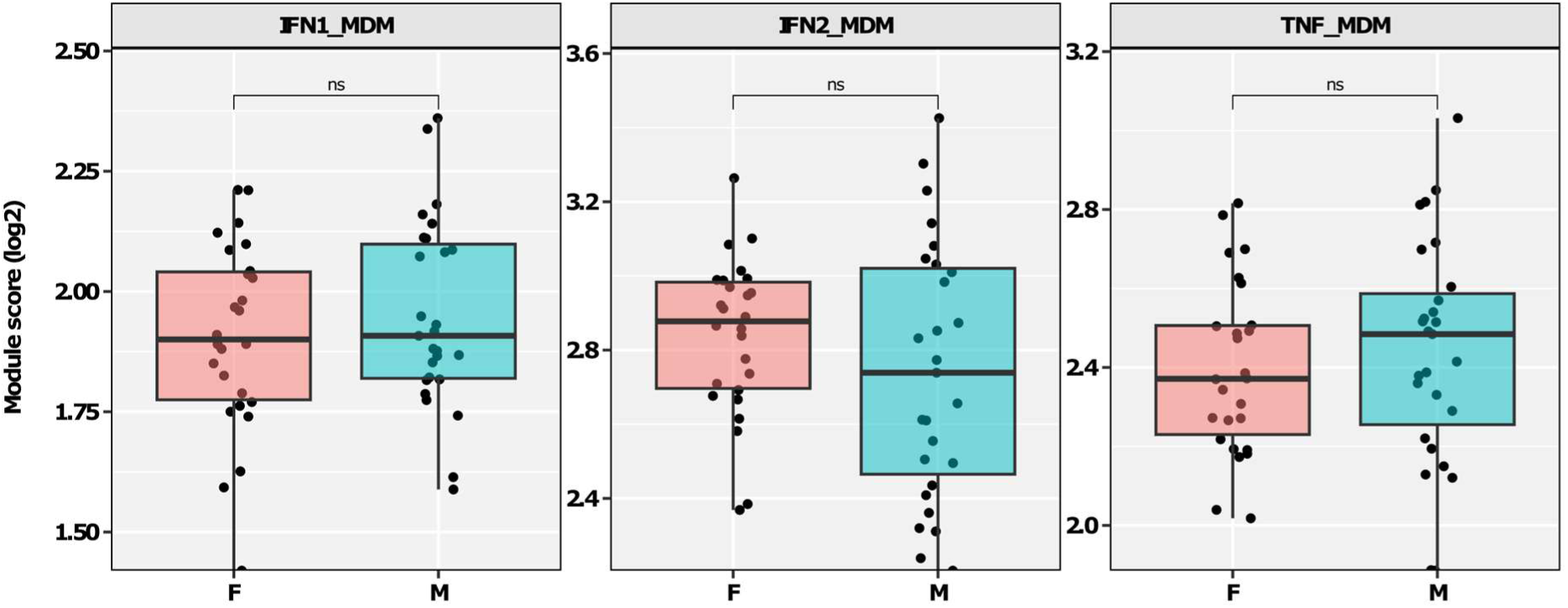
No sex differences in blood immune perturbations during active tuberculosis (TB). Expression of interferon- and TNF-inducible transcriptional modules in bulk RNA sequencing data from whole blood samples of patients with active TB disease (n=27 male, n=26 female). Modules reflect cytokine activity in monocyte-derived macrophages (MDM), derived from stimulant-specific gene expression changes after in vitro MDM stimulation. Boxplots depict median and inter-quartile range and include individual data points. Statistical significance was assessed with unpaired, two-sided Wilcoxon tests (ns p>0.05).

**Supplementary Figure 2.**
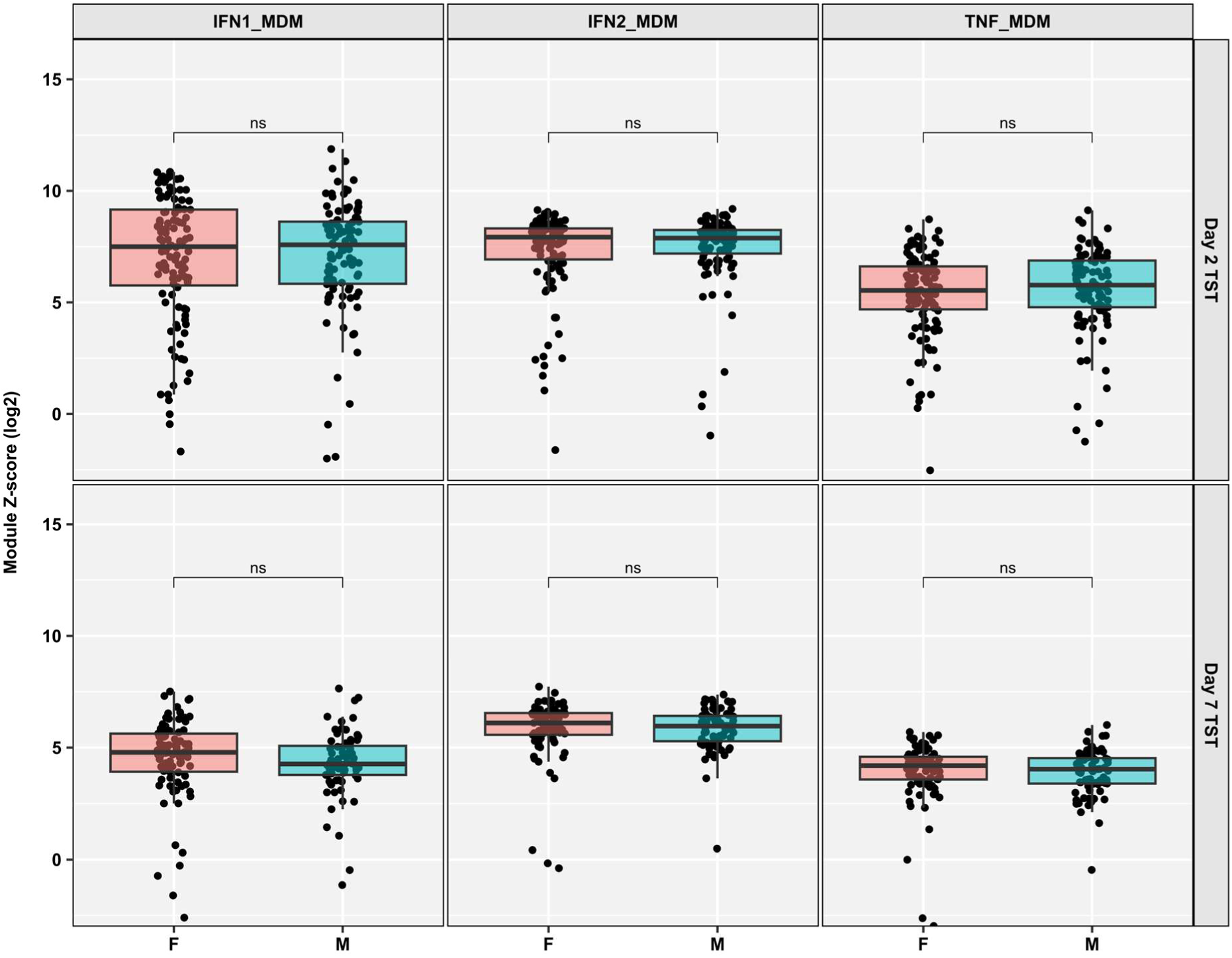
No sex differences in tuberculin skin test (TST) bulk immune responses during latent TB. Expression of interferon- and TNF-inducible transcriptional modules in bulk RNA sequencing data from TST skin biopsies, collected on Day 2 (n=101 male, n=115 female) or Day 7 (n=73 male, n=85 female) following TST challenge. Module scores are shown as Z-score scaled TPM expression using saline-challenged skin biopsies from a separate cohort of participants as control group. Modules reflect cytokine activity in monocyte-derived macrophages (MDM), derived from stimulant-specific gene expression changes after in vitro MDM stimulation. Boxplots depict median and inter-quartile range and include individual data points. Statistical significance was assessed with unpaired, two-sided Wilcoxon tests (ns p>0.05).

**Supplementary Figure 3.**
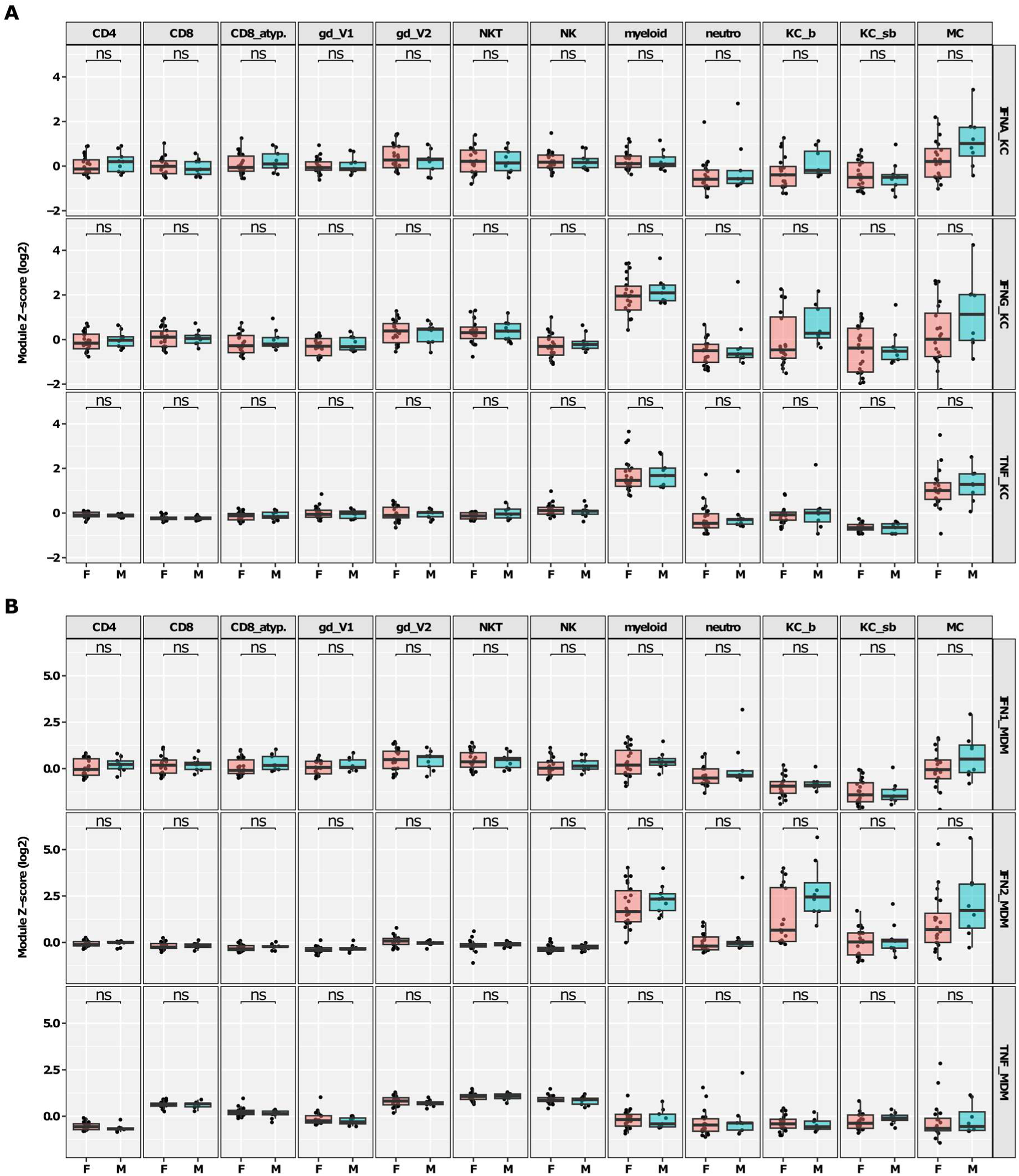
No sex differences in tuberculin skin test (TST) single-cell immune responses during latent TB. Expression of interferon- and TNF-inducible transcriptional modules in single-cell RNA sequencing data from Day 2 TST suction blister cells, shown as average Z-score scaled module score per participant and cell type. Cell types comprise CD4 T cells (CD4), conventional CD8 T cells (CD8), atypical CD8 T cells (CD8_atyp., characterised by reduced expression of CD8B and alpha/beta TCRs compared to conventional CD8s), Vδ1 (gd_V1) and Vδ2 (gd_V2) gamma-delta T cells, natural killer T cells (NKT), natural killer cells (NK), myeloid cells (myeloid), neutrophils (neutro), basal (KC_b) and suprabasal (KC_sb) keratinocytes and melanocytes (MC). Modules reflect cytokine activity in (**A.**) keratinocytes (KC) or (**B.**) monocyte-derived macrophages (MDM), derived from cytokine-specific gene expression changes after in vitro stimulation of KC or MDM. Boxplots depict median and inter-quartile range and include individual data points. Statistical significance between male (M, n=9) and female (F, n=22) samples was assessed with unpaired, two-sided Wilcoxon tests, adjusted for multiple testing (ns FDR>0.05).

